# Differences in domain-specific composition of 24-hour physical behaviors across occupational groups

**DOI:** 10.64898/2026.07.23.26358754

**Authors:** Niko-Petteri Hirvonen, Maisa Niemelä, Anna-Maiju Leinonen, Vahid Farrahi

**Author notes:** Corresponding author: Prof. Dr. Vahid Farrahi, Institute for Sport and Sport Science, Otto-Hahn-Straße 3, 44227 Dortmund, Germany,.

## Abstract

**Purpose:** Physical behaviors, including physical activity, sedentary time, and sleep, are associated with health and together comprise the 24-hour day. Among working-aged adults, the composition of these 24-hour physical behaviors may vary across occupations, but the extent of these differences remains unknown. This population-based study examined how the composition of 24-hour physical behaviors differs in working-aged adults across occupational groups during work and leisure time.

**Methods:** The study population consisted of 2,809 adults from the Northern Finland Birth Cohort 1966 46-year follow-up study, who wore a hip-worn accelerometer for 14 days. The accelerometry data was combined with self-reported diaries to determine time in bed and work hours. Four occupational groups were aggregated based on occupation type and occupational skill level: high-skill white-collar, low-skill white-collar, high-skill blue-collar and low-skill blue-collar. Time spent on physical behaviors during workdays (including working hours and leisure time separately) and non-workdays were calculated. Differences in physical behavior compositions were assessed using a compositional data analysis approach based on log-ratios of geometric means, with the low-skill blue-collar group as the reference group.

**Results:** Significant differences in 24-hour physical behaviors across occupational groups were mainly observed during workdays. During workday working hours, all other occupational groups had less physical activity (both moderate-to-vigorous and light-intensity physical activity) compared to the reference group. During workday leisure time, significant differences were observed in all physical behaviors, except for sedentary time. On non-working days, the high-skill white collar group had higher moderate-to-vigorous physical activity compared to the reference group.

**Conclusions:** The composition of domain-specific 24-hour physical behaviors differs across occupational groups, particularly during workdays.

## Introduction

Physical behaviors comprising the 24-hour day include physical activity, sedentary behavior and sleep, which are collectively associated with numerous health outcomes (1) (2). There has been a conceptual shift from examining the health associations of the time spent in isolation on physical activities, sedentary behavior, and sleep towards considering their combined associations within the 24-hour cycle. However, most studies to date have mainly focused on examining the health associations of 24-hour physical behaviors without considering their domains (3) (4), even though the work and leisure domains in which these behaviors occur may influence their associations with health outcomes (5) (6).

The current physical activity guidelines advocate for an equal amount of physical activity without distinguishing the domain where the behaviors occur (7). For working-aged adults, the occupational domain has a pronounced role in physical behavior accumulation (8) (9). Additionally, occupational physical activity has been proposed to have adverse effects on health outcomes independent of health-enhancing leisure time physical activity (10) (11). Although several studies have indicated existence of the aforementioned physical activity paradox (12) (13), the influence of the occupational type on the composition of 24-hour physical behaviors remains unclear. Only a small number of large-scale studies using a compositional approach have examined separate work and leisure time physical behaviors of working-aged adults through self-reported physical activity (14), self-reported occupation (15) or fixed work schedules (16) highlighting the need for registry-based population-level studies.

Understanding how occupations influence 24-hour physical behaviors is key to informing the development of future guidelines and interventions aimed toward working-aged adults in various occupational contexts. Socioeconomic patterning in physical activity (higher occupational physical activity in lower socioeconomic status vs. higher leisure time physical activity in higher socioeconomic status) is well established in high-income countries (17) (18)

(19). However, the complexity of occupational physical behaviors with potential implications for health outcomes may be overlooked by a simplistic categorization of occupations (i.e. white-collar workers vs. blue-collar workers) (20) (21) (22) (23). Moreover, studies have proposed that work-related health risk factors may cluster in low-skill occupations (24) (22) and that physical behavior patterns may differ in both type and domain by occupational group (25) (26) (27) (19). One population-based study using a compositional approach has suggested that differences may occur in compositional 24-hour physical behaviors across weekdays and weekends by occupational class (28). However, the differences between work and leisure domains within a workday remain unclear.

Currently, device-based assessment of 24-hour physical behaviors is the preferred method of choice in many large-scale observational and epidemiological studies. However, differentiating between work and leisure domains using continuous wearable device data remains challenging. Aside from distinction between weekdays and weekends (28), fixed working schedules have been used to differentiate work and leisure physical behaviors (29). Given that many occupations do not adhere to fixed working schedules, such approaches may not necessarily yield accurate estimations of domain-specific physical behaviors. This methodological challenge can be resolved by complementing wearable-measured data with self-reported work schedules, thereby enabling a more accurate assessment of domain-specific 24-hour physical behaviors.

This study aimed to examine how wearable-measured 24-hour physical behaviors of working-aged adults differ at the population level across work and leisure domains when the type and skill level of the occupation is also considered.

## Materials and methods

The data for this study were drawn from the population-based longitudinal Northern Finland Birth Cohort 1966 (NFBC1966) (30) (31), which comprises all individuals with an expected birth in 1966 (N=12,058) in Northern Finland. The members of the ongoing cohort have been regularly monitored prospectively with broad clinical measurements, interviews, and questionnaires. The participants have given informed consent for participating in the NFBC1966 study and have the right to refuse participation and to withdraw their consent later without providing a reason. This study included cohort members who participated in the follow-up study at the age of 46 (2012-2014, N=5,832). The 46-year follow-up study included questionnaires, attending a clinical examination day, wearing an accelerometer for the measurement of daily physical behaviors, and self-reporting the time in bed and work periods in a paper-based diary. Data access for this study was approved by Statistics Finland (socioeconomic variables) and the Finnish Social and Health Data Permit Authority (Findata). The study was carried out in conformance with the Declaration of Helsinki and was approved by the Ethics Committee of the Northern Ostrobothnia Hospital District (94/2011).

### Measurement of 24-hour physical behaviors

Participants’ waking physical behaviors were measured using a hip-worn accelerometer Hookie AM20 (Traxmeet Ltd., Finland). The participants were instructed to wear the accelerometer during all waking hours for 14 consecutive days, except for water-based activities. Simultaneously to wearing the accelerometer, the participants recorded their wake-up times, work times, and bedtimes in a paper-based diary for the same 14-day measurement period. Information recorded in the diaries was used to determine between a working and a non-working day and to differentiate between physical behaviors during working hours and leisure time.

### Accelerometer data processing

Raw acceleration data were collected at a sampling rate of 100 Hz. The data were segmented into 6-second epochs, and the mean amplitude deviation (MAD) was computed for each segment. The MAD values from the Hookie AM20 and the ActiGraph GTX3 accelerometers have demonstrated excellent agreement (32).

From the 6-second MAD values, the device non-wear time was identified and marked using an approach adapted from a widely validated count-based accelerometer data method (33). Specifically, non-wear time was defined as 90 or more consecutive minutes without movement, allowing for interruptions of up to 30 seconds, provided no additional movement was detected within 30 minutes before or after the interruption. The window size used for handling artifactual acceleration was shortened (from 2 minutes to 30 seconds) as a visual inspection of the signals showed greater performance for shorter intervals with high-frequency raw acceleration data.

The 6-second epochs identified as wear time were further classified as sedentary time (≤ 1.5 METs), light-intensity physical activity (>1.5 and <3.0 METs), moderate-intensity physical activity (>3.0 and <6.0 METs), or vigorous-intensity physical activity (≥6 MET) based on the MAD values.

### Differentiating work and leisure time

Self-reported work and time in bed periods from the diaries were cross-referenced with the accelerometer data. The time in bed was computed based on the diary-reported start and end of bedtime, and the corresponding period was excluded from the accelerometer data. For workdays, the remaining wear time periods were categorized into work-related physical behaviors and leisure time physical behaviors using diary-reported work start and end times.

The total time spent in moderate-to-vigorous physical activity (MVPA), light physical activity (LPA) and sedentary behavior was computed separately for both work and leisure. For non-working days, the total time spent in MVPA, LPA and sedentary behavior was computed for all waking hours. All the computed durations were then summed and divided by the number of corresponding valid days to derive the mean time spent on different physical behaviors during working hours on workdays, during leisure time on workdays, and across non-working days.

### Classification of workdays and non-workdays

A valid day was defined as having at least 10 hours of accelerometer wear time during waking hours, together with complete self-reported information on working hours and time spent in bed (34). The accelerometer signals were visually inspected to ascertain that the accelerometer wear time periods aligned with the self-reported diary-based wake periods. Wear time periods with a systematic misalignment of more than one hour or corrupted data due to device malfunction were excluded from the analysis.

A day was classified as a workday if the participants reported a working period, and as a non-working day if no working period was reported. To be considered a valid workday, the minimum accelerometer wear time during working hours had to be at least 4 hours, or more than 75% of the participant’s average daily working time (35) (36). A minimum of two valid workdays together with two valid non-workdays were required for a participant to be included in the analysis (37). The first day when the participants received the accelerometer was excluded from the analysis.

### Occupational grouping

The participants’ occupations were classified according to the Finnish Classification of Occupations 2010, which corresponds to the International Standard Classification of Occupations (ISCO-08) at the 4-digit unit level, with minimal exceptions. Military occupations were excluded from the analysis due to extensive occupational task heterogeneity. The annually confirmed occupational status matching the year of the measurement period was extracted from a national registry (Statistics Finland) for each participant at a two-digit level (Supplemental Table 1). The major groups were then classified into white-collar (groups 1-4) and blue-collar (groups 6-9). For group 5 (service and sales workers) a two-digit level split was implemented, where sub-group 52 (sales workers) was classified as white-collar and sub-groups 51 (personal service workers), 53 (personal care workers) and 54 (protective services workers) were classified as blue-collar occupations due to markedly higher occupational physical demands (38) (39) (24) (40). Finally, the white and blue-collar groups were further aggregated by occupational skill level as high-skill white-collar occupations (major groups 1-3), high-skill blue-collar occupations (major groups 6-7 and sub-groups 51,53,54), low-skill white-collar occupations (major group 4 and sub-group 52), and low-skill blue-collar occupations (major groups 8-9).

### Demographics and work characteristics

Participants’ educational attainment was obtained from the national register maintained by Statistics Finland. Higher education was defined as International Standard Classification of Education (ISCED) levels 5–8. Marital status and household median income were self-reported. Physically strenuous work and job strain (low job control and high occupational demands) were defined using composite scores from two Likert-scale nine-point questionnaires reported in more detail elsewhere (41) (42). Specifically, the participants’ work was considered physically strenuous if they reported above-median composite scores. Job strain was calculated by dividing the mean scores of job demand- and job control items. Participants had to report their work being mentally demanding at least half of the time for their work to be considered mentally demanding.

### Statistical analysis

Participant characteristics were described using standard descriptive statistics. The components of 24-hour physical behaviors are essentially compositional and add up to a total sum (i.e. 24 hours). Any exchange of time among the components of 24-hour physical behaviors requires an equivalent amount from one or multiple other behaviors (43). The properties of time-use data require analytical approaches that acknowledge mutual codependency (44), such as compositional data analysis, which is a methodology capable of respecting the relative nature of time-use data (45).

Following a previous study demonstrating better suitability of compositional data analysis over a standard statistical approach (MANOVA) for examining group differences in time-use data, a compositional data analysis was used (46). To create a 24-hour time-use composition for each participant, the duration of all physical behaviors was linearly rescaled to sum to 24 hours (1,440 minutes) per day. The composition of 24-hour behaviors in each of the four occupational groups was described using compositional means, noting that compositional data analysis cannot include zero-values. In our sample, only one participant had zero-values in MVPA, and was excluded from the analysis.

The differences in daily time-use across the four groups were examined with separate time-use compositions created for workdays and non-workdays. A workday was defined as 7-part composition, consisting of physical behaviors during working hours (MVPA, LPA, and sedentary time) and during leisure time (MVPA, LPA, sedentary time, and time in bed). A non-workday was defined as a 4-part composition (MVPA, LPA, sedentary time, and time in bed). As such, the physical behaviors of both workdays and non-workdays add up to 24 hours within each composition.

Group difference analyses were performed in accordance with published methods for compositional data analysis applicable to time-use data (46). Log-ratios of geometric means of the occupational groups were computed considering the low-skill blue-collar group as a reference. The log-ratios express group differences on a relative scale where a negative value indicates less time spent on behavior, and a positive value indicates more time spent on behavior compared to the reference group. A bootstrap resampling with 1,000 replicates of the same sample size was used to achieve 95% confidence intervals for the group log-ratio difference estimates. The data was analyzed using R 4.4.1. (R Core team, Vienna, Austria).

## Results

A flowchart of the participant inclusion and exclusion is shown in Figure 1. Overall, 54% of those who participated in the clinical examination day and agreed to wear an accelerometer were included in the analytical sample. The main reasons for participant exclusion included missing diary or occupational information, not working during the measurement period, or no recorded accelerometry data for the minimum of 2 workdays and 2 non-workdays. The final analysis consisted of 2,809 participants.

**Figure 1.**
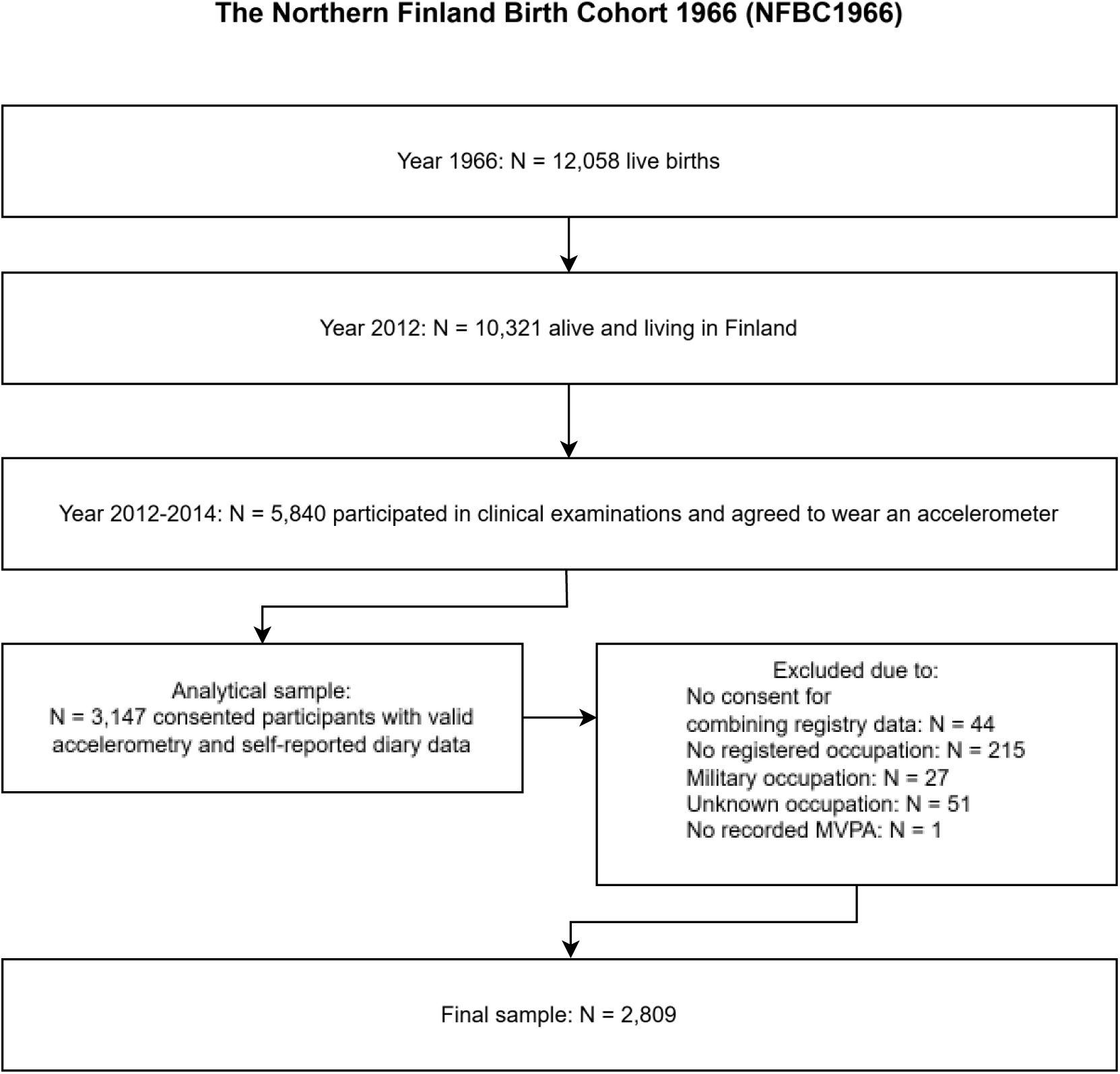
Flowchart of the participant inclusion and exclusion criteria

The participant characteristics are shown in Table 1. Over half of the study population were female (59.9%), married (61.2%) and highly educated (58.9%). The low-skill blue-collar group consisted mostly of men (26.2% female), had the lowest prevalence of mentally challenging work (10.5%), and reported the most physically strenuous work (mean score 6.18, SD 2.53). A high education was most prevalent in the high-skill white-collar group (85.9%) in conjunction with highest reported household gross income (75,000, IQR 49,985). The low-skill white-collar group reported the highest job strain (mean score 1.29, SD 0.34).

**Table 1.** Participant characteristics by occupational group.

|  | Low-skill<br>blue-collar<br>workers<br>(N = 507) | High-skill<br>blue-collar<br>workers<br>(N = 419) | Low-skill<br>white-collar<br>workers<br>(N = 336) | High-skill<br>white-collar<br>workers<br>(N = 1,547) | All<br>(N = 2,809) |
| --- | --- | --- | --- | --- | --- |
| **Demographics** |  |  |  |  |  |
| Female | 26.2% | 81.9% | 78.9% | 60.8% | 59.9% |
| Missing (%) | 0 (0%) | 0 (0%) | 0 (0%) | 0 (0%) | 0 (0%) |
| Married | 53.1% | 56.3% | 55.1% | 66.5% | 61.2% |
| Missing (%) | 22 (4%) | 16 (4%) | 12 (4%) | 44 (3%) | 94 (3%) |
| High education | 12.8% | 14.4% | 51.1% | 85.9% | 58.9% |
| Missing (%) | 53 (12%) | 24 (6%) | 29 (9%) | 33 (2%) | 139 (5%) |
| Household gross income,<br>median €/p.a., (IQR) | 52,000<br>(28,000) | 48,000<br>(33,500) | 55,000<br>(44,000) | 75,000<br>(49,985) | 62,000<br>(45,000) |
| Missing (%) | 74 (15%) | 56 (13%) | 35 (10%) | 122 (8%) | 287 (10%) |
| **Work characteristics** |  |  |  |  |  |
| Mentally challenging work | 10.5% | 19.3% | 53.9% | 76.3% | 53.3% |
| Missing (%) | 54 (11%) | 28 (7%) | 15 (5%) | 67 (4%) | 164 (6%) |
| Physically strenuous<br>work, mean score (SD) | 6.18 (2.53) | 5.95 (2.35) | 4.06 (2.89) | 2.50 (2.37) | 3.83 (2.95) |
| Missing (%) | 68 (13%) | 43 (10%) | 34 (10%) | 114 (7%) | 254 (%) |
| Job strain, mean score (SD) | 1.21 (0.34) | 1.26 (0.30) | 1.28 (0.34) | 1.16 (0.28) | 1.20 (0.30) |
| Missing (%) | 55 (12%) | 28 (7%) | 15 (5%) | 69 (5%) | 162 (%) |

The accelerometer-derived information of the study population is shown in Table 2. On average, the participants had 7.6 (SD 2.3) valid workdays and 4.7 (SD 2.0) valid non-workdays. Overall, all occupational groups had similar wear time across workdays and non-workdays.

**Table 2.**
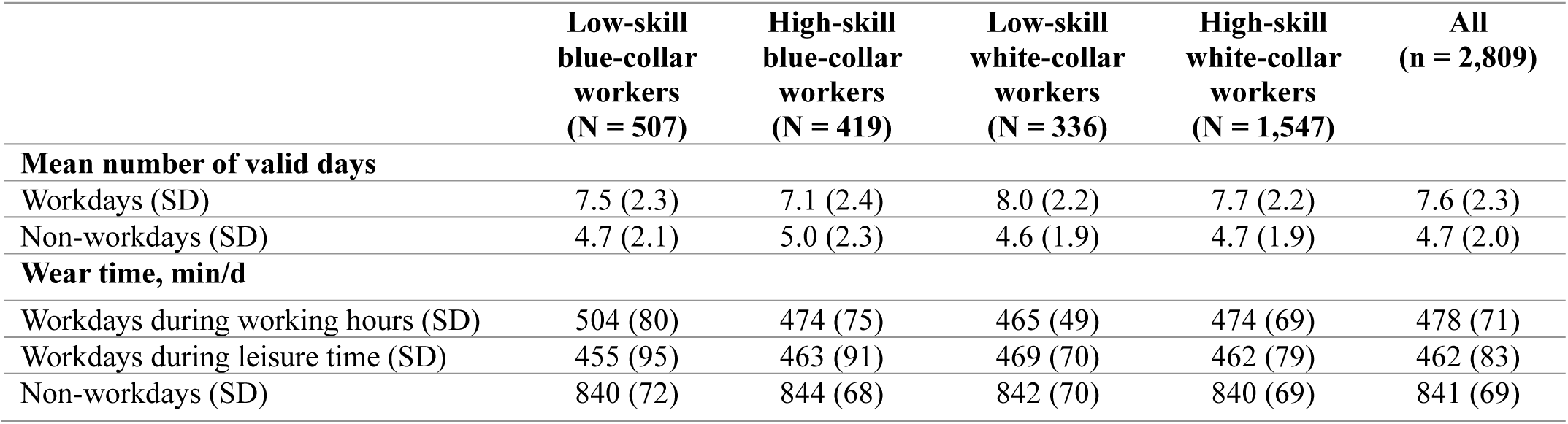
Accelerometer-assessed information of the study population.

|  | Low-skill<br>blue-collar<br>workers<br>(N = 507) | High-skill<br>blue-collar<br>workers<br>(N = 419) | Low-skill<br>white-collar<br>workers<br>(N = 336) | High-skill<br>white-collar<br>workers<br>(N = 1,547) | All<br>(n = 2,809) |
| --- | --- | --- | --- | --- | --- |
| **Mean number of valid days** |  |  |  |  |  |
| Workdays (SD) | 7.5 (2.3) | 7.1 (2.4) | 8.0 (2.2) | 7.7 (2.2) | 7.6 (2.3) |
| Non-workdays (SD) | 4.7 (2.1) | 5.0 (2.3) | 4.6 (1.9) | 4.7 (1.9) | 4.7 (2.0) |
| **Wear time, min/d** |  |  |  |  |  |
| Workdays during working hours (SD) | 504 (80) | 474 (75) | 465 (49) | 474 (69) | 478 (71) |
| Workdays during leisure time (SD) | 455 (95) | 463 (91) | 469 (70) | 462 (79) | 462 (83) |
| Non-workdays (SD) | 840 (72) | 844 (68) | 842 (70) | 840 (69) | 841 (69) |

The compositional domain-specific time-use by occupation group is shown in Table 3. Across all groups, sedentary time accounted for the largest proportion of waking hours during workdays and non-workdays. Sedentary time was also the most prevalent behavior during working hours across all groups, ranging from 53% of the time spent working for the low-skill blue-collar group to 78% of time spent working for the high-skill white-collar group. Both low-skill and high-skill blue-collar groups accrued more MVPA and LPA during working hours compared to workday leisure time, whereas both low-skill and high-skill white-collar groups accrued more MVPA and LPA during workday leisure time.

**Table 3.**
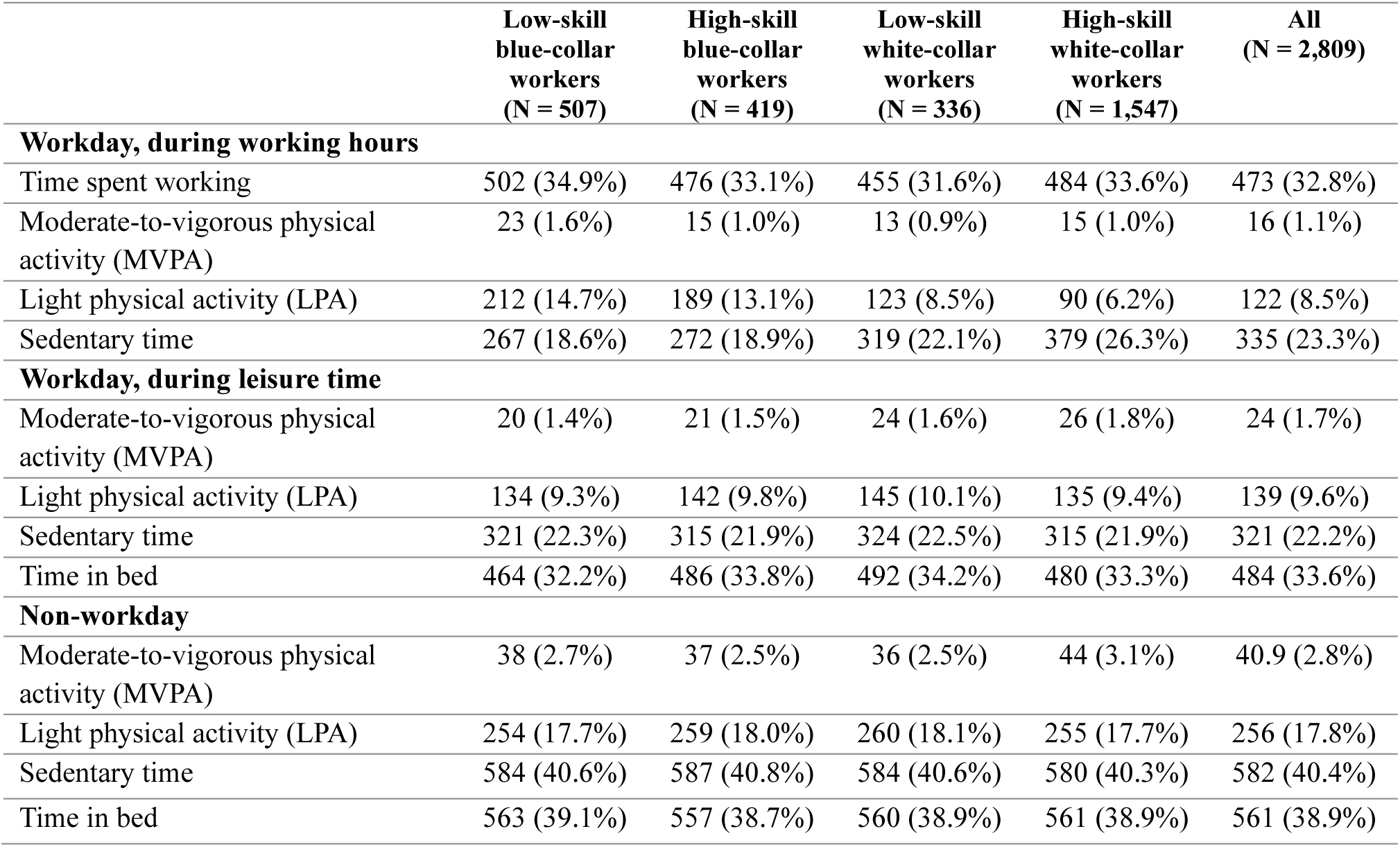
Domain-specific group mean time-use compositions. The values are minutes/day (% of 24-hour day) linearly scaled to add up to 1440 minutes.

|  | Low-skill<br>blue-collar<br>workers<br>(N = 507) | High-skill<br>blue-collar<br>workers<br>(N = 419) | Low-skill<br>white-collar<br>workers<br>(N = 336) | High-skill<br>white-collar<br>workers<br>(N = 1,547) | All<br>(N = 2,809) |
| --- | --- | --- | --- | --- | --- |
| **Workday, during working hours** |  |  |  |  |  |
| Time spent working | 502 (34.9%) | 476 (33.1%) | 455 (31.6%) | 484 (33.6%) | 473 (32.8%) |
| Moderate-to-vigorous physical activity (MVPA) | 23 (1.6%) | 15 (1.0%) | 13 (0.9%) | 15 (1.0%) | 16 (1.1%) |
| Light physical activity (LPA) | 212 (14.7%) | 189 (13.1%) | 123 (8.5%) | 90 (6.2%) | 122 (8.5%) |
| Sedentary time | 267 (18.6%) | 272 (18.9%) | 319 (22.1%) | 379 (26.3%) | 335 (23.3%) |
| **Workday, during leisure time** |  |  |  |  |  |
| Moderate-to-vigorous physical activity (MVPA) | 20 (1.4%) | 21 (1.5%) | 24 (1.6%) | 26 (1.8%) | 24 (1.7%) |
| Light physical activity (LPA) | 134 (9.3%) | 142 (9.8%) | 145 (10.1%) | 135 (9.4%) | 139 (9.6%) |
| Sedentary time | 321 (22.3%) | 315 (21.9%) | 324 (22.5%) | 315 (21.9%) | 321 (22.2%) |
| Time in bed | 464 (32.2%) | 486 (33.8%) | 492 (34.2%) | 480 (33.3%) | 484 (33.6%) |
| **Non-workday** |  |  |  |  |  |
| Moderate-to-vigorous physical activity (MVPA) | 38 (2.7%) | 37 (2.5%) | 36 (2.5%) | 44 (3.1%) | 40.9 (2.8%) |
| Light physical activity (LPA) | 254 (17.7%) | 259 (18.0%) | 260 (18.1%) | 255 (17.7%) | 256 (17.8%) |
| Sedentary time | 584 (40.6%) | 587 (40.8%) | 584 (40.6%) | 580 (40.3%) | 582 (40.4%) |
| Time in bed | 563 (39.1%) | 557 (38.7%) | 560 (38.9%) | 561 (38.9%) | 561 (38.9%) |

MVPA was higher during workdays compared to non-workdays for both the low-skill blue-collar group (43 min vs. 38 min) and the low-skill white-collar group (37 min vs. 36 min). The high-skill blue-collar group had an equal amount sedentary time on workdays and non-workdays, whereas all other groups had less sedentary time on workdays compared to non-workdays. All groups spent less time in bed on workdays compared to non-workdays.

The mean differences in 24-hour physical behaviors across occupational groups during workday working hours and workdays leisure time are presented in Figure 2a and Figure 2b, respectively. Compared to the low-skill blue-collar group, all other groups engaged in less MVPA during working hours (-0.41, 95% confidence interval (CI) = -0.53 to -0.29; -0.54, 95% CI = -0.67 to -0.41; and -0.40, 95% CI = -0.49 to -0.33 in the high-skill blue-collar, low-skill white-collar and the high-skill white-collar group, respectively). Similarly, all other groups engaged in less LPA during working hours compared to the reference group (-0.09, 95% CI -0.16 to -0.02; -0.52, 95% CI -0.61 to -0.44; -0.83, 95% CI -0.88 to -0.79 in the high-skill blue-collar, low-skill white-collar and the high-skill white-collar group, respectively). More sedentary time during working hours compared to the reference group was observed in the low-skill white-collar group (0.19, 95% CI = 0.12 to 0.26) and in the high-skill white-collar group (0.37, 95% CI = 0.33 to 0.42).

**Figure 2.**
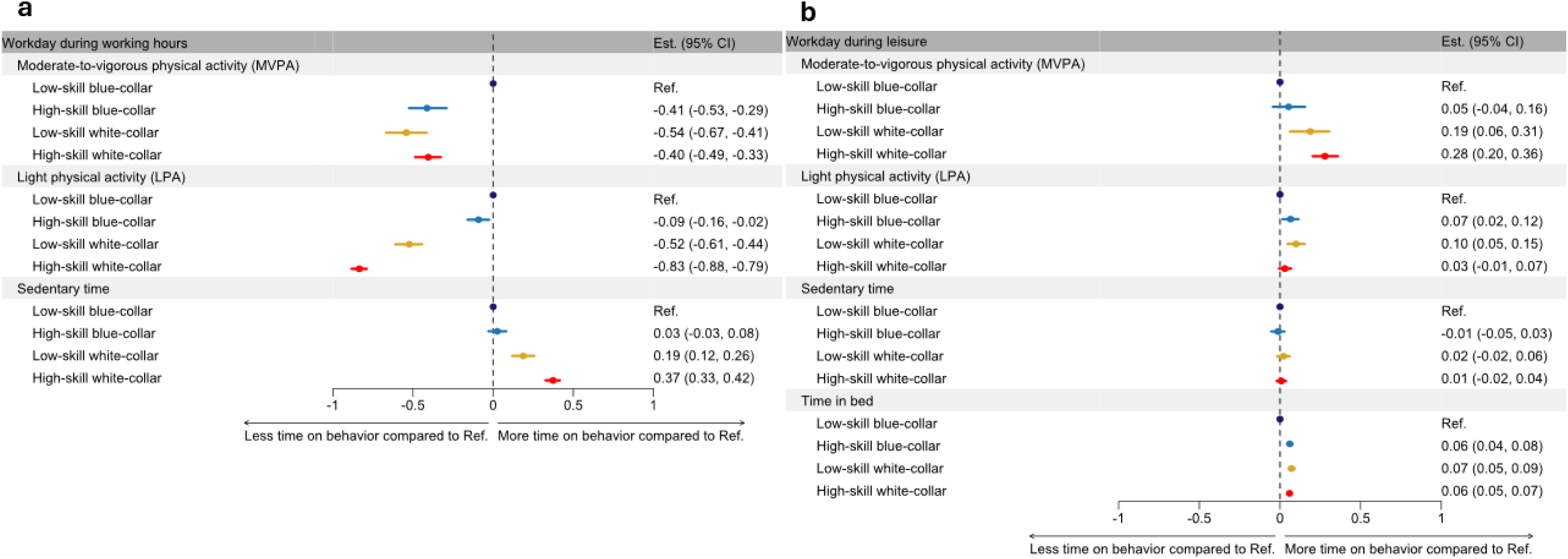
Grouped forest plot of log-ratio differences on workdays during working hours (a) and during leisure time (b). The low-skill blue-collar was the reference group. If the 95% confidence interval contains 0, no significant difference between the two groups for the behavior is considered.

During workday leisure time, both white-collar groups engaged in more MVPA (0.19, 95% CI = 0.06 to 0.36; 0.28, 95% CI = 0.20 to 0.36 in the low-skill white-collar group, and the high-skill white-collar group, respectively) compared to the low-skill blue-collar group. More LPA during workday leisure time was observed for the high-skill blue-collar group (0.07, 95% CI = 0.02 to 0.12) and the low-skill white-collar group (0.10, 95% CI = 0.05 to 0.15) compared to the reference group. No statistically significant differences between the groups in terms of sedentary time spent during workday leisure time compared to the reference group were observed. All other groups had more time in bed on workdays than the reference group.

Mean differences in 24-hour physical behaviors across occupational groups on non-workdays are presented in Figure 3. On non-workdays, the high-skill white-collar group had more MVPA (0.14, 95% CI = 0.05 to 0.22) compared to the low-skill blue-collar group. No other statistically significant differences compared to the reference group were observed.

**Figure 3.**
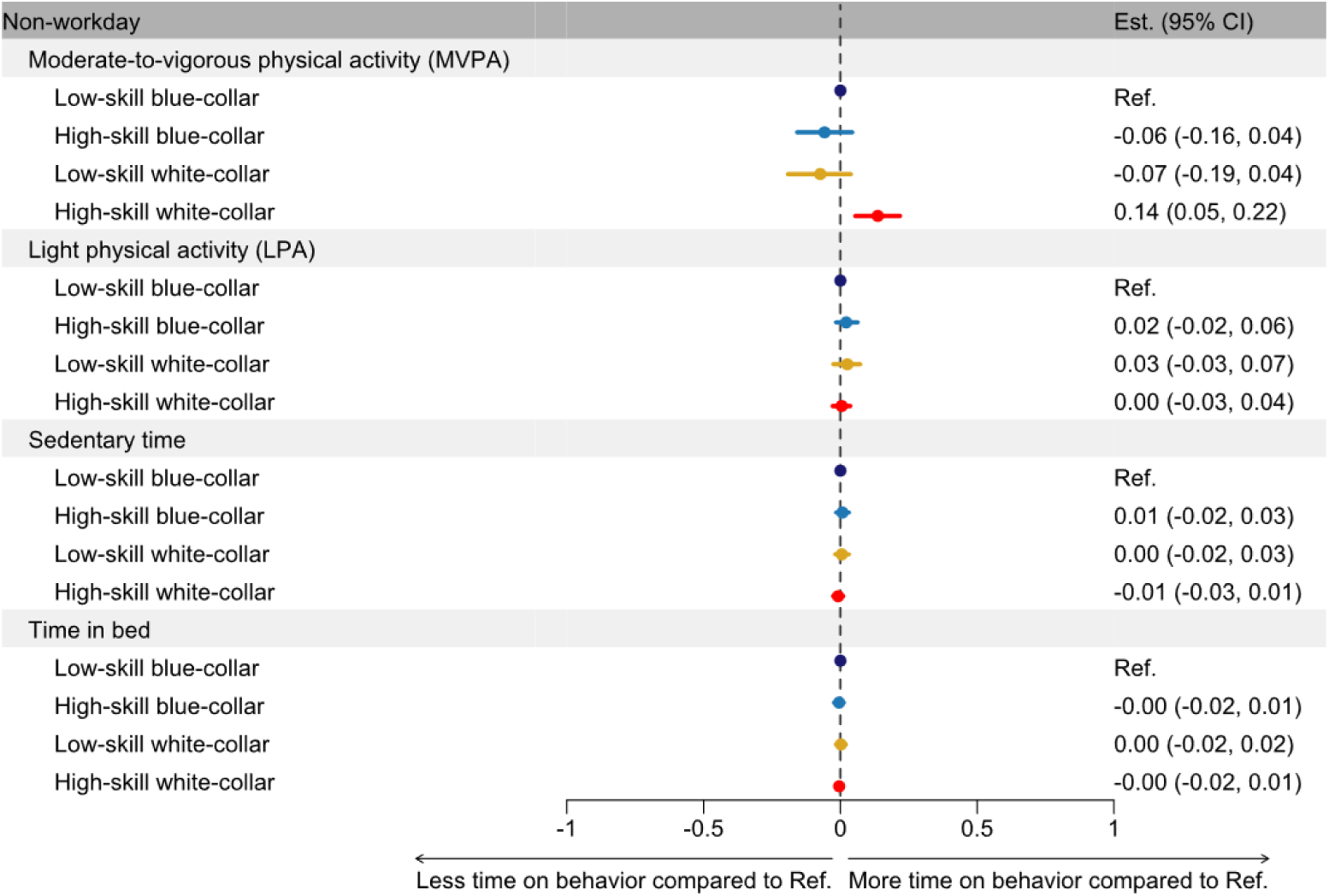
Forest plot of log-ratio differences during non-workdays. The low-skill blue-collar group was the reference group. If the 95% confidence interval contains 0, no significant difference between the two groups for the behavior is considered.

## Discussion

This population-based study examined differences in domain-specific physical behaviors of working-aged adults across occupational groups using a compositional data analysis approach. Most apparent differences in the composition of 24-hour physical behaviors were observed on workdays during working hours. During workday leisure time, the differences in physical behaviors were most apparent for MVPA. The only significant difference on non-workdays was higher MVPA engagement in the high-skill white-collar group compared to the low-skill blue-collar group (reference group). Collectively, these findings suggest that differences in 24-hour physical behaviors among working-aged adults may be mainly attributable to workdays.

This study is the first to describe differences across the full spectrum of 24-hour physical behaviors separately during working hours, leisure time on workdays, and non-workdays in working-aged adults. Earlier studies investigating how occupations influence physical behaviors separately during work and leisure have been mainly performed with smaller numbers of participants including workers from blue-collar (14) (15) (46) (47) or white-collar occupations (16) (29). These studies have consistently shown that differences in working-aged adults’ physical behavior profiles are associated with occupational type. Our findings extend the body of evidence by demonstrating that these differences could be primarily attributable to working hours on workdays, whereas there could be relatively fewer differences in leisure time and non-workday physical behaviors.

Higher physical activity during working hours, especially MVPA, seems to be inversely related to physical activity during workday leisure time. Compared to the low-skill blue-collar group, all other occupational groups accumulated relatively less physical activity (both MVPA and LPA) during working hours but more physical activity during workday leisure time. Previous studies have reported relatively unhealthier physical activity profiles for workers with high occupational physical activity (48) (49), making the observed differences between work and leisure during workdays particularly noteworthy. The lower physical activity levels observed during working hours are likely explained primarily by differences in job type and demands (39) (50) (51). Our study findings further suggest that occupational differences may also differentially influence physical behaviors during leisure time on workdays.

All occupational groups engaged in higher leisure time physical activity on workdays (both MVPA and LPA) compared to the reference group. Occupational factors such as long working hours (11), high job strain (42) and the need for recovery from work (52) (53) may influence personal resource thresholds towards engaging in health-enhancing leisure time physical activity. Notably, the occupational groups with higher self-reported physically strenuous work had lower levels of leisure-time MVPA. In addition, both high-skill groups across white and blue-collar workers engaged in relatively more MVPA during their leisure time on a workday. This finding contrasts with a previous study using a compositional data analysis approach which showed that high-skill workers engaged in lower weekday MVPA compared to low-skill workers (28). Our findings further highlight that the low-skill blue-collar workers mainly accrue their workday MVPA during working hours whereas all other occupational groups accumulate relatively more MVPA during workday leisure time.

Differences in sedentary time during working hours were mainly observed across the white-collar groups. However, sedentary time during workday leisure time was comparable across all occupational groups. Previous studies examining domain-specific sedentary time suggest differential effects for sedentary time during work and leisure (54) (55). More work-related sedentary time for workers in physically demanding occupations could potentially increase participation in health-enhancing leisure time physical activity through, for example, reduced fatigue (56) (57). In contrast, additional sedentary time for workers in desk-based occupations has been suggested to be detrimental to health (58). It is plausible to assume that sedentary time during working hours across white-collar groups may be subject to occupational skill level. Skill-related occupational characteristics, such as greater job autonomy (nested within self-reported job strain), may enable high-skill white-collar workers to accumulate more sustained sedentary time by allowing longer periods of uninterrupted work (59) (60).

The only significant difference on non-workdays was that high-skill white-collar workers engaged in more MVPA, compared to the reference group. These results are consistent with previous studies reporting less differences in physical behaviors during non-workdays between occupational groups (21) (28) (60). Although the differences in other components of physical behaviors were non-significant, there may be a complex holistic interrelationship between physical behavior and its’ component parts on workdays and non-workdays (61). For example, the high amount of MVPA of the low-skill blue-collar group during working hours may be compensated through less leisure time physical activity during workday leisure time, but not on non-workdays (11) (62) (63). Further studies are needed to understand how such complex interrelationships and differences in physical behaviors are related to different health outcomes across occupational groups.

## Strengths and limitations

The strengths of this study include the relatively large population-based sample of working-aged adults. The occupational groups were formed considering both job type and occupational skill level. The combined accelerometry and self-reported diary data methodology fills a research gap by separately describing working hours, workday leisure time, and days off work, which was previously unavailable at the population level. Previous studies have typically monitored physical behaviors over a 7-day measurement period. The 14-day measurement protocol (with 7.6 and 4.7 average valid workdays and non-workdays, respectively) is another strength of this study because it provides a more realistic and accurate estimates of habitual physical behaviors.

This study is not without limitations. The study was cross-sectional in design, and therefore temporal or causal relationships cannot be inferred. Physical behavior properties (e.g. bout durations) were not differentiated, limiting our scope to the total time accumulated on each behavior. Although wearable hip-worn accelerometry has been shown to provide a good estimate of the intensity of movement, the hip-worn devices could possibly underestimate the intensity of upper-extremity dominant physical behaviors. Additionally, the group comparisons were based on group averages in absolute intensity, which might not reflect the actual physiological load. The time spent in bed and working periods was recorded using self-reported diaries, which are subject to recall bias. Furthermore, the study participants were more likely to be female, married, have children, and have a higher socio-economic status compared to the non-participants. This may have introduced a degree of selection bias and may influence the generalizability of the findings (31).

## Conclusions

In conclusion, the results of this study highlight a job type effect on physical behavior profiles in working-aged adults across various occupational skill levels. The composition of 24-hour physical behaviors across occupational groups differ mainly within the work and leisure domains on workdays. On non-working days, the composition of physical behaviors among adults appears to be similar across occupational groups, with the only differences observed in MVPA. These findings suggest that physical activity guidelines for working-aged adults may need to account for the influence of occupational demands on 24-hour physical behaviors.

## Acknowledgements

We wish to thank all cohort members, researchers and NFBC project center personnel who participated in the NFBC1966 data collections.

## Confllict of Interest

The authors declare no conflict of interest. The results of the study are presented clearly, honestly, and without fabrication, falsification, or inappropriate data manipulation.

## Data Availability Statement

NFBC data are available from the University of Oulu, Infrastructure for Population Studies. Permission to use the data can be applied for research purposes via an electronic material request portal. In the use of data, we follow the EU general data protection regulation (679/2016) and the Finnish Data Protection Act.

The use of personal data is based on a cohort participant’s written informed consent in their latest follow-up study, which may cause limitations to its use.

Please, contact the NFBC project center (NFBCprojectcenter(at)oulu.fi) and visit the cohort website (www.oulu.fi/nfbc) for more information.

## Appendices

**Supplemental Table 1.**
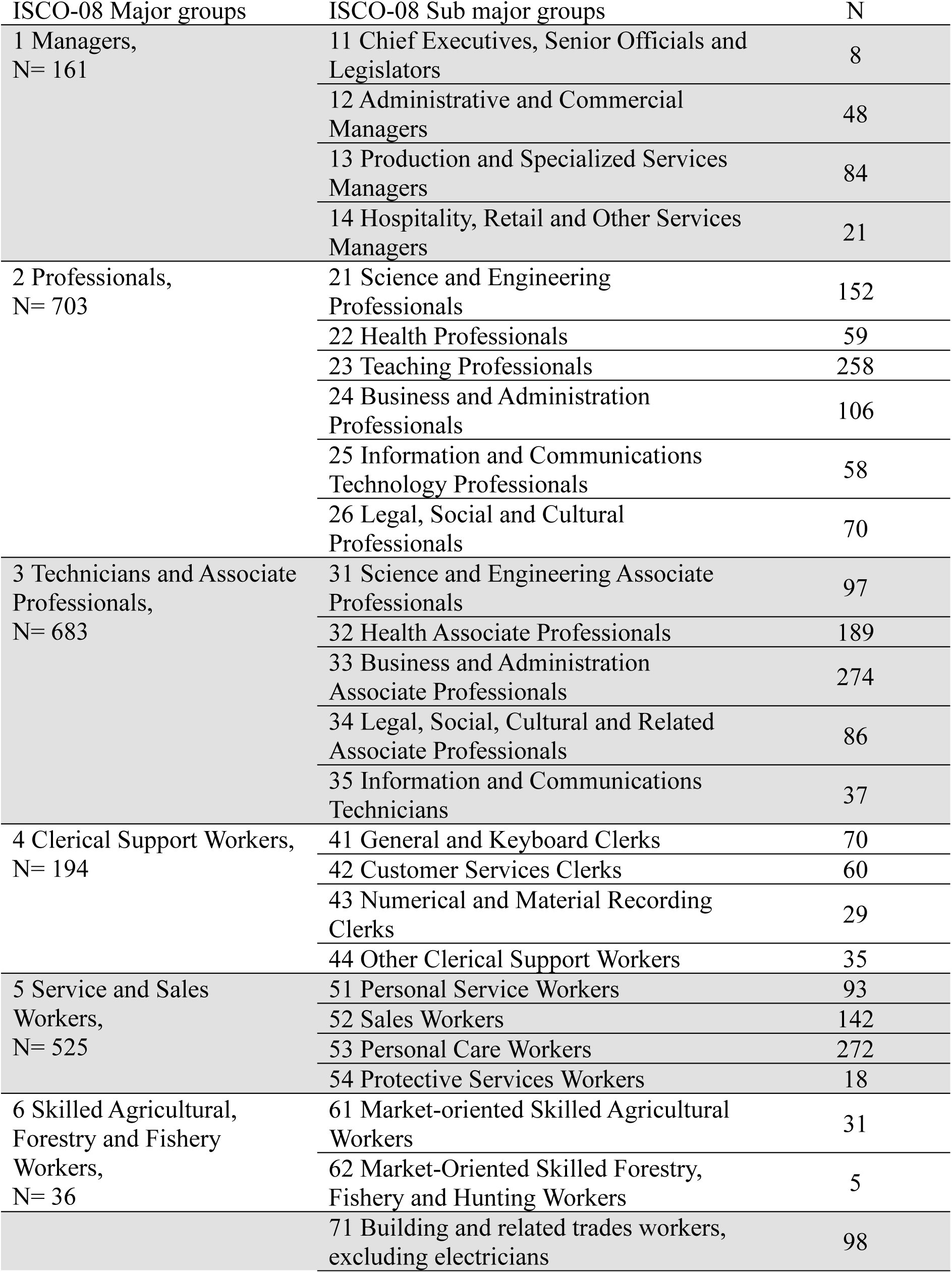
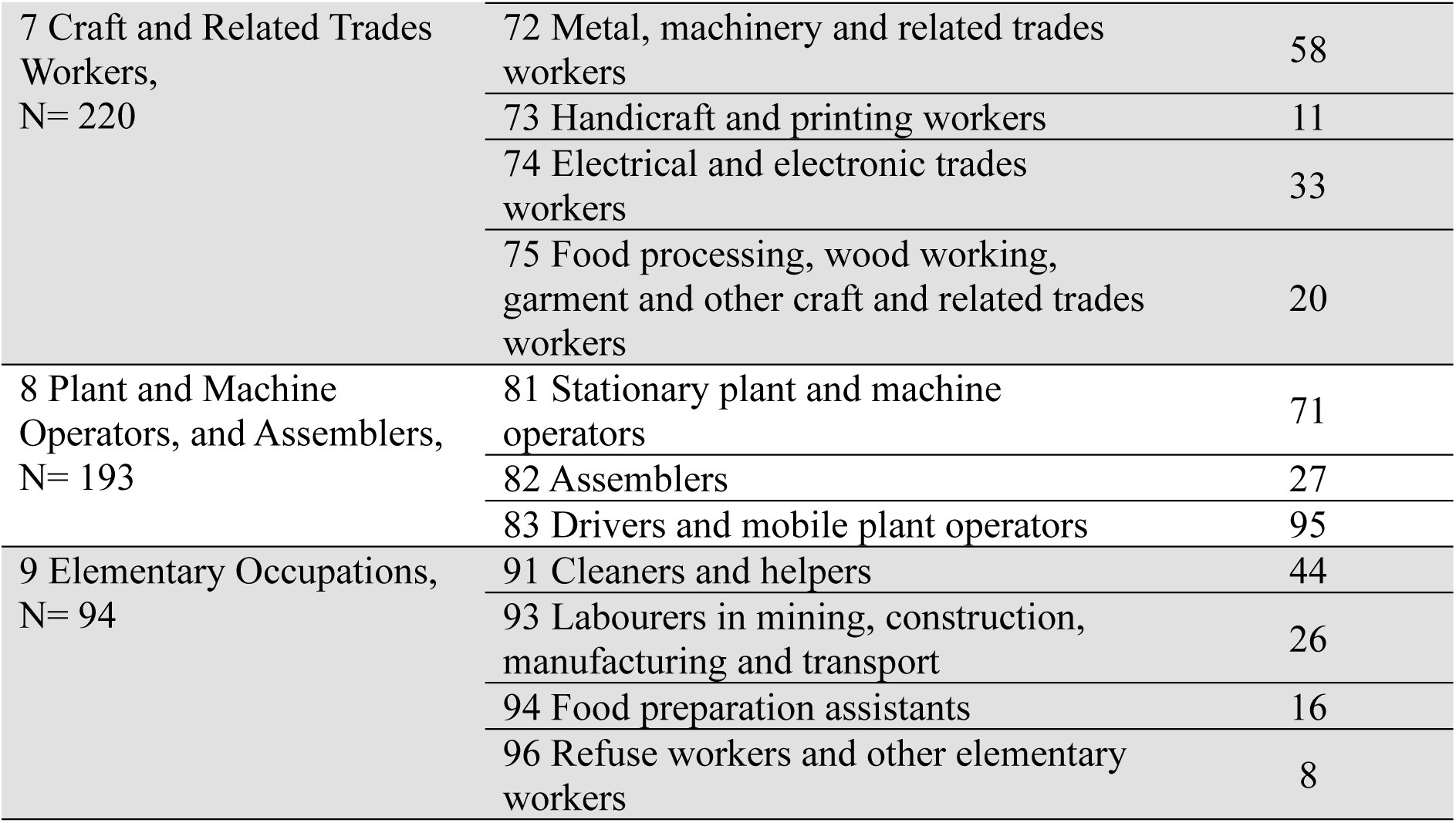
Distribution of study sample participants across ISCO-08 major- and sub major groups.

| ISCO-08 Major groups | ISCO-08 Sub major groups | N |
| --- | --- | --- |
| 1 Managers,<br>N= 161 | 11 Chief Executives, Senior Officials and Legislators | 8 |
|  | 12 Administrative and Commercial Managers | 48 |
|  | 13 Production and Specialized Services Managers | 84 |
|  | 14 Hospitality, Retail and Other Services Managers | 21 |
| 2 Professionals,<br>N= 703 | 21 Science and Engineering Professionals | 152 |
|  | 22 Health Professionals | 59 |
|  | 23 Teaching Professionals | 258 |
|  | 24 Business and Administration Professionals | 106 |
|  | 25 Information and Communications Technology Professionals | 58 |
|  | 26 Legal, Social and Cultural Professionals | 70 |
| 3 Technicians and Associate Professionals,<br>N= 683 | 31 Science and Engineering Associate Professionals | 97 |
|  | 32 Health Associate Professionals | 189 |
|  | 33 Business and Administration Associate Professionals | 274 |
|  | 34 Legal, Social, Cultural and Related Associate Professionals | 86 |
|  | 35 Information and Communications Technicians | 37 |
| 4 Clerical Support Workers,<br>N= 194 | 41 General and Keyboard Clerks | 70 |
|  | 42 Customer Services Clerks | 60 |
|  | 43 Numerical and Material Recording Clerks | 29 |
|  | 44 Other Clerical Support Workers | 35 |
| 5 Service and Sales Workers,<br>N= 525 | 51 Personal Service Workers | 93 |
|  | 52 Sales Workers | 142 |
|  | 53 Personal Care Workers | 272 |
|  | 54 Protective Services Workers | 18 |
| 6 Skilled Agricultural, Forestry and Fishery Workers,<br>N= 36 | 61 Market-oriented Skilled Agricultural Workers | 31 |
|  | 62 Market-Oriented Skilled Forestry, Fishery and Hunting Workers | 5 |
|  | 71 Building and related trades workers, excluding electricians | 98 |
| 7 Craft and Related Trades Workers,<br>N= 220 | 72 Metal, machinery and related trades workers | 58 |
|  | 73 Handicraft and printing workers | 11 |
|  | 74 Electrical and electronic trades workers | 33 |
|  | 75 Food processing, wood working, garment and other craft and related trades workers | 20 |
| 8 Plant and Machine Operators, and Assemblers,<br>N= 193 | 81 Stationary plant and machine operators | 71 |
|  | 82 Assemblers | 27 |
|  | 83 Drivers and mobile plant operators | 95 |
| 9 Elementary Occupations,<br>N= 94 | 91 Cleaners and helpers | 44 |
|  | 93 Labourers in mining, construction, manufacturing and transport | 26 |
|  | 94 Food preparation assistants | 16 |
|  | 96 Refuse workers and other elementary workers | 8 |

